# Performance of ICD-10-based injury severity scores in pediatric trauma patients using the ICD-AIS map and survival rate ratios

**DOI:** 10.1101/2023.12.04.23299239

**Authors:** Rayan Hojeij, Pia Brensing, Michael Nonnemacher, Bernd Kowall, Ursula Felderhoff-Müser, Marcel Dudda, Christian Dohna-Schwake, Andreas Stang, Nora Bruns

**Affiliations:** Department of Pediatrics I, Neonatology, Pediatric Intensive Care Medicine, and Pediatric Neurology, University Hospital Essen, University of Duisburg-Essen, Essen, Germany; TNBS, Centre for Translational Neuro- and Behavioural Sciences, University Hospital Essen, University of Duisburg-Essen, Essen, Germany; Institute for Medical Informatics, Biometry and Epidemiology, University Hospital Essen, Essen, Germany; Department of Trauma, Hand and Reconstructive Surgery, University Hospital Essen, Essen, Germany

**Keywords:** Injury severity, International Classification of diseases, Mortality, Pediatric trauma, survival probabilities

## Abstract

**Objective:** Our study aimed to identify the superior predictor of mortality from International Classification of Diseases 10 (ICD-10) codes among pediatric trauma patients in the German hospital database (GHD), a nationwide database comprising all hospitalizations in the country.

**Study design and setting:** Hospital admissions of patients aged < 18 years with injury-related ICD-10 codes were selected. The maximum abbreviated injury scale (MAIS) and injury severity score (ISS) were calculated using the ICD-AIS map provided by the Association for the Advancement of Automotive Medicine, which we adjusted to the German modification of the ICD-10 classification. The survival risk ratio was used to calculate the single worst ICD-derived injury (single ICISS) and a multiplicative injury severity score (multiplicative ICISS). The ability to predict mortality of the four above mentioned scores were compared in the selected trauma population and within four clinically relevant subgroups using discrimination and calibration.

**Results:** Out of 13,992,596 cases < 18 years of age hospitalized between 2014 and 2020, 1,720,802 were trauma patients and ICD-AIS mapping was possible in 1,328,377 cases. Mortality was highest in patients with only one coded injury. Cases with mapping failure (n = 392,425; 22.8 %) were younger and had a higher mortality rate. SRR-derived scores had a better discrimination calibration than ICD-AIS based scores in the overall cohort and all four subgroups (AUC ranges between 0.985 and 0.998 versus 0.886 and 0.972 respectively).

**Conclusion:** Empirically derived measures of injury severity were superior to ICD-AIS mapped scores in the GHD to predict mortality in pediatric trauma patients. Given the high percentage of mapping failure and high mortality among cases with single coded injury, the single ICISS may be the most suitable measure of injury severity in this group of patients.

## 1. Introduction

Traumatic injuries in children and adolescents are a serious public health concern and among the leading causes of mortality and acquired morbidity [1, 2]. Every year, pediatric injuries account for almost one million deaths worldwide [3, 4]. Mortality is associated with the severity of the injury in trauma patients, making its measurement an important cornerstone of risk assessment [5, 6]. Precise risk assessment is further needed for clinical evaluation, selection of the appropriate treatment center, and clinical decision support in the pre- and intrahospital phase. At the same time, it is used for the assessment of trauma center performance, benchmarking, and research purposes [7-9].

The injury severity score (ISS), which is based on the Abbreviated Injury Scale (AIS), is one of the most widely used scoring systems for adult and pediatric trauma patients[10-12]. This ISS score is based on the severity of injuries in specific anatomical regions, each classified according to the AIS classification. The ISS is then calculated from the AIS scores as the sum of the squares of the highest AIS grade in each of the three most severely injured areas [13, 14]. AIS scores are assigned by experts based on clinical information according to the *Abbreviated Injury Scale (AIS) 2005* manual [15]. If immediate clinical information is unavailable, for example in datasets using routine health care data, AIS scores can be derived from international classification of disease (ICD) codes to calculate the ISS [16-28].

An alternative approach to assess injury severity from ICD codes are empirical measures that can be calculated directly from the dataset of interest based on the survival risk ratio (SRR). The SRR reflects the probability of surviving a specific diagnosis compared to all other individuals in the same dataset. Subsequently, the single worst injury or a multiplicative injury severity score (ICISS) can be derived. In trauma research, ICISS approaches have been successfully applied in large administrative health care datasets to predict in-hospital mortality in the absence of clinical information on injury severity [29-33]. The use of administrative datasets in Germany is currently evolving. These datasets are of special interest in the field of pediatric trauma and pediatric critical care research to overcome small case numbers in single or multicenter observational studies [34].

Despite the broad application of ICD-AIS and SRR-based methods in adult and pediatric trauma patients, clinically important subpopulations of pediatric trauma patients are not assessed such as traumatic brain injury. Further, these measures have not been established for pediatric trauma patients in the German hospital database (GHD). This database is unique, as it contains all hospitalizations to public hospitals of the entire country. Because no private children’s hospitals exist in Germany, the GHD represents the total number of pediatric hospitalizations.

The aim of this study was to compare the performances of the ICD-AIS and SRR-based injury severity scores using this dataset to predict mortality in pediatric trauma patients and clinically relevant subpopulations. Accordingly, the best identified method by this comparison will be used for risk adjustment for future pediatric trauma research in the GHD.

## 2. Methods

### 2.1. Study design and database

This study is a retrospective study comparing the performance of different injury severity scores for the pediatric trauma patients in the German hospital dataset (GHD). The reimbursement for public German hospitals is based on diagnosis related groups (DRG). Hospitals are required to transmit data on every hospitalization to the Hospital Remuneration System (InEK) by law (§21 KHEntgG). As the transmission of data is a prerequisite for reimbursement, hospitals have a strong incentive to supply comprehensive data sets. At the InEK, data are checked for plausibility, anonymized, and forwarded to the Federal Statistical Office (FSO), where the GHD is hosted.

Access to the GHD is granted to researchers upon written request. Information on the structure of the GHD is provided by the FSO and has been described in detail previously. Further information is available at https://www.forschungsdatenzentrum.de/en/health/drg.

### 2.2. Data extraction and study population

Cases of patients < 18 years of age treated in public German hospitals and discharged between 2014 and 2020 were eligible. We extracted cases with an injury as main diagnosis at discharge according to the International Classification of Diseases, 10th Edition, German Modification (ICD-10-GM) (ICD “S” and “T” codes from chapter XIX). Based on the ICD-coded injury pattern, the following subgroups of patients were defined:

- TBI main diagnosis: Traumatic brain injury coded as the main diagnosis (ICD-HD).
- TBI secondary diagnosis: Traumatic brain injury has been coded as a secondary diagnosis with another injury as main diagnosis.
- Multiple injuries: At least two injuries were coded as main and secondary diagnosis, not TBI.
- Single injury: patients with only one injury coded.

Survival to discharge was extracted from the discharge reason, which is mandatory for each case.

### 2.3. Abbreviated Injury Scale, Injury Severity Score, and ICD-AIS mapping

AIS scores are assigned according to the anatomical region and type of injury [15]. The severity of each injury is classified on an ordinal scale ranging from 1 (minor injury) to 6 (untreatable or non-survivable). A score of 9 is assigned to injuries of unclassified or unclassifiable severity. Each injury is assigned to one of the nine AIS chapters representing different body regions. The maximum AIS (MAIS) is the highest single AIS score assigned to one of the 9 AIS chapters.

The ISS is used to quantify composite injury severity in patients with multiple injuries. It grades the severity of injury according to the AIS codes but assigns these grades to only six body regions. Possible ISS scores range from 1 to 75 and are calculated from the sum of the squared values of the three most severely injured body regions. In case one injury is coded “6” (= unsurvivable injury), the overall score is set to the maximum score of 75; if one region is classified as 9, the ISS cannot be calculated [12]. See **Error! Reference source not found**. for differences between AIS and ISS scoring systems.

To derive AIS and ISS scores from ICD-10 codes, we purchased the expert-developed ICD-AIS map 2005 (update 2008) from the AAAM (Association for the Advancement of Automotive Medicine) [22]. The ICD-AIS map is based on the ICD-10 clinical modification (CM), whereas the GHD uses the German modification (GM) of the ICD-10 system. For this reason, we assessed the ICD-AIS map 2005 (update 2008) for incompatibilities between the ICD-10 systems and actively searched for codes that are present in the GM but not in the CM. Incompatibilities and missing codes were replaced according to Niemann et al. [28, 34, 35]. If more than one AIS grade was applicable, the lower/lowest possible AIS grade was selected.

To calculate the AIS values for each AIS chapter and ISS body region, the GHD was transformed into longitudinal form and merged with the ICD-AIS map. After retransformation into the original format, the MAIS and the ISS were extracted and calculated for each case.

### 2.4. Survival risk ratios and ICISS

The International Classification of Disease Injury Severity Score (ICISS) is an empirical ICD-based method derived from so-called ‘survival risk ratios’ (SRR) to predict mortality after injury [36]. SRRs are calculated for each ICD-10 code in the data set by dividing the number of cases with a given ICD-10 code who survived by the total number of cases with this ICD-10 code in the dataset. The highest possible value is 1.0, with lower values representing higher lethality [29].

We calculated the single worst injury score (Single-ICISS) by extracting the lowest SRR for each case. Additionally, we calculated a multiplicative injury score (Multiple-ICISS) by multiplying all SRRs values of a case [37, 38].

### 2.5. Missing Data

There were no missing data on age, main diagnoses, and survival at discharge. Missing data on secondary diagnoses were impossible to detect because we could not determine whether the diagnosis was not present or not coded.

### 2.6. Data analysis

All calculations and analyses were performed using SAS release 9.4 and SAS Enterprise Guide 7.3 (SAS Institute, Cary, North Carolina, USA). Logistic regressions were used to examine in-hospital mortality as the main outcome, using the four computed injury severity scores as predictor variables. Cases where ICD-AIS mapping was not possible were removed from performance analyses. Thus, the model’s discrimination of each score was tested on the entire cohort of trauma patients in whom ICD-AIS mapping could be performed (Fig. 1**Error! Reference source not found**.) and the four subgroups by calculating receiver operating curves, areas under the curve (AUC) with 95 % confidence intervals (CI). AUC, also known as c-statistics or concordance statistics, is the proportion of concordant pairs among all possible pairs made of one survivor and one non-survivor. The calibration assessment was conducted using the Hosmer-Lemeshow (H-L) chi-squared statistics, with a lower value indicating better calibration. The assessment of statistical significance for the H-L test, typically gauged through p-values, with p<0.1 indicates poor calibration. However, the sensitivity of the H-L test increases with larger samples, potentially yielding statistically significant results that may not necessarily reflect practical significance [39]. In our study, we did not assess H-L p-values due to the very large sample size. Consequently, we assessed supplementary measures of model fit by computing the slope and intercept of the calibration curves. The best model calibration will have a slope near one and the intercept close to zero [40].

**Fig. 1.**
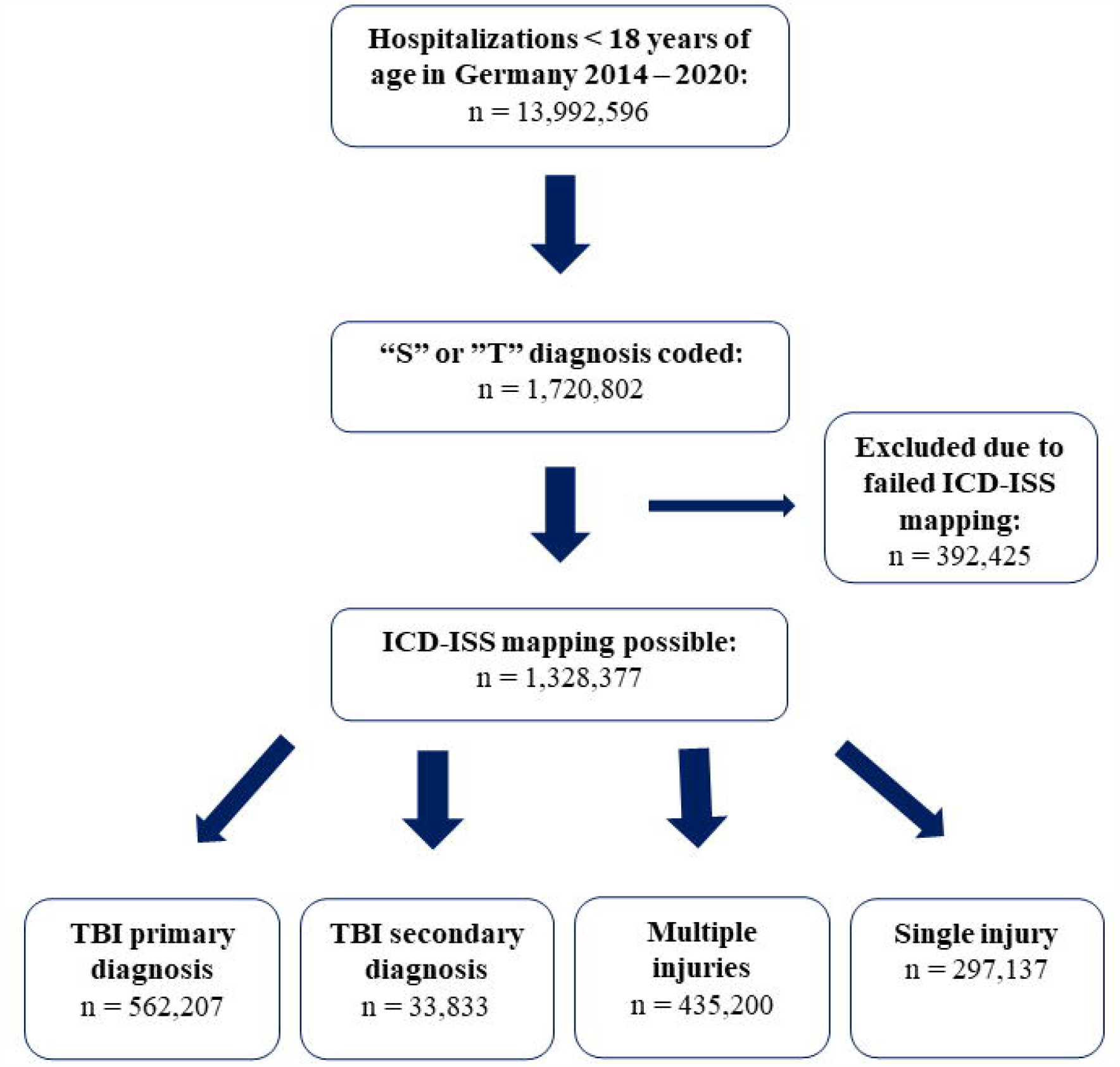
Flowchart of included injury cases aged 0-17 years in Germany, 2014-2020. Abbreviations : ICD=International classification of Diseases 10^th^ edition, ISS= Injury severity Score, TBI = Traumatic Brain Injury. “S” or “T” diagnosis codes=ICD-10 codes of chapter XIX including codes starting with either S or T.

## 3. Results

Between 2014 and 2020, 13,992,596 hospitalizations in patients < 18 years of age occurred in Germany. Of these, 1,720,802 million records had a main diagnosis code eligible for ICD-AIS mapping (fig 1). 77.2 % (n = 1,328,377) of eligible cases could be assigned a valid AIS score for all AIS chapters, with assigned scores ranging from 1 to 6. The remaining cases either received an AIS severity score of 9 due to an unclassifiable injury according to the AIS system or due to insufficient information in the description of the ICD-10-GM code.

In total, 22% of cases were lost for performance analysis due to ICD-AIS mapping failure. Mapping failure was more frequent in the youngest (49.4%) compared to the oldest age group (21.3%) (Table 1).

**Table 1.** Descriptive statistics based on the possibility to apply ICD-AIS mapping among pediatric injury cases in Germany, 2014-2020.

|  | Possible ICD-AIS map<br>(N= 1,328,377) | Failed ICD-AIS map<br>(N= 392,425) |
| --- | --- | --- |
|  | n (%) | n (%) |
| Age (years) |  |  |
| 0-3 | 351,154 (26.4) | 193,649(49.3) |
| 4-6 | 188,851 (14.2) | 47,045 (12.0) |
| 7-12 | 375,736 (28.3) | 68,313 (17.4) |
| 13-17 | 412,636 (31.1) | 83,418 (21.3) |
| Sex |  |  |
| Female | 539,598 (40.6) | 179,568 (45.8) |
| Male | 788,779 (59.4) | 212,857 (54.2) |
| Subgroups |  |  |
| TBI main diagnosis | 562,207 (42.3) | 1,783 (0.45) |
| TBI secondary diagnosis | 33,833 (2.55) | 1,236 (0.32) |
| Multiple injuries | 435,200 (32.76) | 54,818 (13.97) |
| Single injury | 297,137 (22.37) | 334,588 (85.26) |
| Deceased | 863 (0.07) | 244 (0.06) |

Abbreviations : ICD: International classification of Diseases 10<sup>th</sup> edition, ISS: Injury Severity Score, TBI: Traumatic brain injury.

In cases with failed ICD-ISS map, 0.45% have TBI main diagnosis, and 0.32% have TBI secondary diagnosis, whereas the highest proportion of unmappable codes (85.26%) were observed in the single injury subgroup. Among deceased cases with mapping failure, 62.7 % belonged to the single injury group, while in the multiple injury and TBI main diagnosis groups 16.4 % and 17.2 % could not be mapped, respectively (Table 2).

**Table 2.** Numbers and percentages of in-hospital death among male and female injured children or adolescents in Germany with respect to ICD-AIS mapping, 2014-2020.

|  | Possible ICD-AIS map |  | Failed ICD-AIS map |  |
| --- | --- | --- | --- | --- |
|  | Survived<br>(n = 1,327,514)<br>n(%) | Deceased<br>(n = 863)<br>n(%) | Survived<br>(n = 392,181)<br>n(%) | Deceased<br>(n = 244)<br>n(%) |
| TBI main diagnosis | 561,784 (42.29) | 423 (49.01) | 1,743 (0.44) | 40 (16.39) |
| TBI secondary diagnosis | 33,747 (2.54) | 86 (9.96) | 1,227 (0.31) | 9 (3.69) |
| Multiple injuries | 434,996 (32.75) | 204 (23.64) | 54,776 (13.97) | 42 (17.21) |
| Single injury | 296,987 (22.36) | 150 (17.38) | 334,435 (85.28) | 153 (62.70) |

Abbreviations : ICD: International classification of Diseases 10<sup>th</sup> edition, ISS: Injury Severity Score, TBI: Traumatic brain injury

MAIS and ISS scores were markedly lower in survivors than in non-survivors in the overall cohort and subgroups (Table 3).

**Table 3.** Descriptive statistics of severity measurements among survivors and non-survivors of injured children or adolescents in Germany stratified by subgroups, 2014-2020.

|  |  | Surviving<br>(n=1,327,514) | Non-surviving<br>(n=863) |
| --- | --- | --- | --- |
|  |  | Median<br>(10<sup>th</sup> – 90<sup>th</sup> percentile) | Median<br>(10<sup>th</sup> – 90<sup>th</sup> percentile) |
| MAIS | Total population | 1 (1-2) | 3 (3-3) |
|  | TBI main diagnosis | 1 (1-1) | 3 (3-3) |
|  | TBI secondary diagnosis | 2 (1-3) | 3 (3-4) |
|  | Multiple injuries | 2 (1-2) | 3 (2-4) |
|  | Single injury | 2 (1-2) | 3 (3-3) |
| ISS | Total population | 1 (1-4) | 10 (9-27) |
|  | TBI main diagnosis | 1 (1-2) | 17 (9-27) |
|  | TBI secondary diagnosis | 4 (1-10) | 18 (9-34) |
|  | Multiple injuries | 4 (1-5) | 10 (4-25) |
|  | Single injury | 4 (1-4) | 9 (9-9) |
| ICISS-Single | Total population | 0.999 (0.995-0.999) | 0.554 (0-0.789) |
|  | TBI main diagnosis | 0.999 (0.995-0.999) | 0.554 (0-0.769) |
|  | TBI secondary diagnosis | 0.997 (0.954-0.999) | 0.521 (0-0.789) |
|  | Multiple injuries | 0.999 (0.992-0.999) | 0.392 (0-0.789) |
|  | Single injury | 0.999 (0.995-1) | 0.515 (0-0.944) |
| ICISS-Multiple | Total population | 0.999 (0.992-0.999) | 0.082 (0-0.5444) |
|  | TBI main diagnosis | 0.999 (0.995-0.999) | 0.134 (0-0.474) |
|  | TBI secondary diagnosis | 0.995 (0.931-0.999) | 0.044 (0-0.501) |
|  | Multiple injuries | 0.998 (0.987-0.999) | 0.048 (0-0.536) |
|  | Single injury | 0.999(0.995-1) | 0.075 (0-0.921) |

Abbreviations : MAIS: maximum score on the Abbreviated Injury Scale, ISS: Injury Severity Score, ICISS: ICD-derived multiple injury severity score, TBI: Traumatic Brain Injury

Accordingly, single and multiplicative ICISS were lower in non-survivors. The highest ICISS derived scores were observed in the single injury group, where half of the deceased patients were not analyzed due to ICD-AIS mapping failure (Fig. 2).

**Fig. 2.**
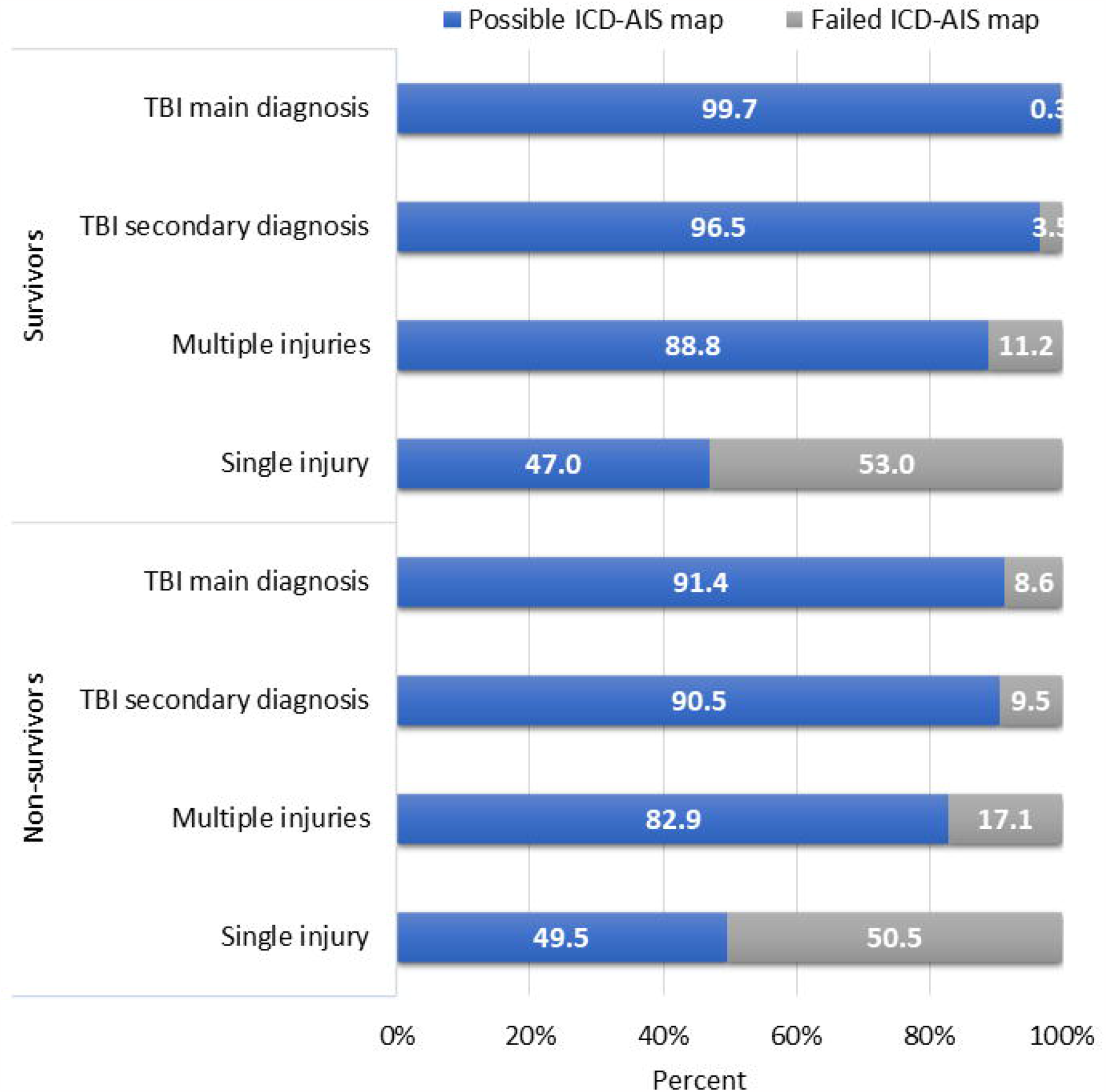
Percentages of ICD-AIS mapping among subgroups stratified by survival status. Abbreviations : ICD=International classification of Diseases 10^th^ edition, ISS= Injury severity Score, TBI = Traumatic Brain Injury.

### 3.1. Model discrimination

Discrimination between survivors and non-survivors was excellent for all injury scores in the overall cohort and subgroups with a minimum AUC value of 0.89 (Table 4).

**Table 4.** Models’ discrimination and Hosmer and Lemeshow calibration statistics of severity measurements by population type of injured children and adolescents in Germany.

|  | ICD-AIS mapping |  |  |  | SRR based scores |  |  |  |
| --- | --- | --- | --- | --- | --- | --- | --- | --- |
|  | MAIS |  | ISS |  | ICISS-Single |  | ICISS-Multiple |  |
|  | AUC<br>95% CI | H-L | AUC<br>95% CI | H-L | AUC<br>95% CI | H-L | AUC<br>95% CI | H-L |
| Total population<br>(n=1,328,377) | 0.942<br>(0.932-0.951) | 758.05 | 0.951<br>(0.941-0.961) | 592.61 | 0.994<br>(0.990-0.998) | 121.47 | 0.995<br>(0.972-0.999) | 53.32 |
| TBI main diagnosis<br>(n=562,207) | 0.968<br>(0.955-0.981) | 293.11 | 0.972<br>(0.959-0.985) | 251.95 | 0.995<br>(0.989-0.999) | 47.73 | 0.995<br>(0.999-1) | 22.07 |
| TBI secondary<br>diagnosis<br>(n=33,833) | 0.903<br>(0.877-0.929) | 93.79 | 0.931<br>(0.902-0.961) | 37.89 | 0.997<br>(0.995-0.999) | 30.83 | 0.997<br>(0.996-0.999) | 2.42 |
| Multiple injuries<br>(n=435,200) | 0.886<br>(0.858-0.914) | 270.72 | 0.904<br>(0.876-0.933) | 167.77 | 0.998<br>(0.997-0.999) | 27.99 | 0.998<br>(0.997-0.999) | 9.16 |
| Single injury<br>(n=297,137) | 0.939<br>(0.915-0.963) | 134.07 | 0.939<br>(0.915-0.963) | 207.46 | 0.986<br>(0.972-1) | 35.83 | 0.985<br>(0.970-1) | 19.79 |

Abbreviations: SRR: Survival Risk ratios, MAIS: Maximum Abbreviated Injury Score, ISS: Injury Severity Score, ICISS: International classification of Diseases 10<sup>th</sup> Edition based Injury Severity Score, AUC: Area under the curve, H-L: Hosmer and Lemeshow chi square , 95%CI : 95% confidence interval, TBI: Traumatic Brain Injury.

We observed the highest AUCs among the empirically derived ICISS scores, with AUCs better than 0.99.

### 3.2. Model calibration

Considerable differences of H-L chi square test statistics were observed between ICD-AIS mapped scored and empirically derived injury scores, with H-L statistics ranging from 2.42 to 758.05 (Table 4). The highest values of the H-L statistical were observed for the MAIS and ISS injury scores models (i.e., higher H-L statistics, indicating poorer model calibration). Overall, the ICISS models performed best, both on the full population and the four subgroups. Slope and intercept showed very well-calibrated models, with the multiplicative ICISS displaying the slope closest to 1 and intercept nearest to zero for the full cohort and all four subgroups, which confirmed the superiority of the multiplicative ICISS in the H-L test statistics (Table 5).

**Table 5.** Calibration slope and intercept of calibration curves of severity measurements by population type of injured children and adolescents in Germany.

|  | ICD-AIS mapping |  |  |  | SRR based scores |  |  |  |
| --- | --- | --- | --- | --- | --- | --- | --- | --- |
|  | MAIS |  | ISS |  | ICISS-Single |  | ICISS-Multiple |  |
|  | Slope | Intercept | Slope | Intercept | Slope | Intercept | Slope | Intercept |
| Total population<br>(n=1,328,377) | 1.58<br>(0.98;2.18) | -0.000<br>(-0.002;0.002) | 1.80<br>(1.20;2.40) | -0.002<br>(-0.001;0.000) | 1.16<br>(1.16;1.17) | -0.000<br>(-0.000;-0.000) | 1.07<br>(1.07;1.08) | -0.000<br>(-0.000;-0.000) |
| TBI main diagnosis<br>(n=562,207) | 1.08<br>(-1.50;3.52) | 0.002<br>(-0.02;0.03) | 1.37<br>(0.25;2.50) | 0.002<br>(-0.005;0.007) | 1.14<br>(1.13;1.14) | -0.000<br>(-0.000;-0.000) | 1.07<br>(1.06-1.07) | -0.000<br>(-0.000;-0.000) |
| TBI secondary<br>diagnosis<br>(n=33,833) | 1.12<br>(-0.68; 2.93) | 0.001<br>(-0.008;0.011) | 1.32<br>(1.00;1.65) | -0.000<br>(-0.002;0.001) | 1.09<br>(1.08;1.09) | -0.000<br>(-0.000;-0.000) | 1.03<br>(1.02;1.03) | -0.000<br>(-0.000;-0.000) |
| Multiple injuries<br>(n=435,200) | 1.47<br>(0.43;2.51) | 0.000<br>(-0.001;0.001) | 2.06<br>(1.21-2.91) | -0.000<br>(-0.001;0.000) | 1.15<br>(1.15-1.16) | -0.000<br>(-0.000;-0.000) | 1.05<br>(1.04-1.05) | -0.000<br>(-0.000;-0.000) |
| Single injury<br>(n=297,137) | 1.88<br>(1.44;2.32) | -0.000<br>(-0.001;0.000) | 2.68<br>(2.02;3.33) | -0.000<br>(-0.001;0.000) | 1.28<br>(1.27;1.29) | -0.000<br>(-0.000;-0.000) | 1.18<br>(1.17;1.19) | -0.000<br>(-0.000;-0.000) |

Abbreviations: SRR: Survival Risk ratios, ICD: International classification of Diseases 10<sup>th</sup> Edition, MAIS: Maximum Abbreviated Injury Score, ISS: Injury Severity Score, ICISS: International classification of Diseases 10<sup>th</sup> Edition based Injury Severity Score, TBI: Traumatic Brain Injury.

## 4. Discussion

This study assessed ICD-AIS- and SRR-based methods to predict mortality in pediatric trauma patients in the GHD and found excellent discrimination for both methods. Mapping between ICD and ISS codes failed in over 20% of cases, and these cases exhibited disparities in terms of age, mortality, and the coded injury patterns when compared to successfully mapped cases. Model performances in clinically relevant subpopulations were separately analyzed and yielded similar results as in the overall cohort.

The youngest age group exhibited a higher percentage of unmappable ICD codes compared to older cases. This discrepancy may be caused by the circumstance that very mild injuries frequently have unspecific and thus unmappable codes. Age-specific differences in injury patterns are widely acknowledged, with adolescents experiencing high impact trauma with severe and ICD-AIS mappable injuries, more frequently than infants and young children [41]. Another relevant observation is the frequent failure of ICD-AIS mapping in cases with only one coded injury, which may also result from unspecific codes in mildly injured patients. At the same time, mortality was highest in the single injury group, suggesting that patients may have died in the emergency room before comprehensive injury assessment had been performed. The single injury group may therefore be a heterogenous group of the least and most severely injured patients in the dataset. However, the clinically and epidemiologically most relevant type of pediatric trauma is traumatic brain injury. Mapping failure was below 0.5 % in cases with TBI as main or secondary diagnosis, reflecting the importance of the subgroup analyses we conducted.

The discriminatory ability to predict in-hospital deaths was outstanding in all four assessed scoring systems and for all subgroups, with an AUC ranging from 0.89 to 0.99. Within this excellent performance, the ICISS-based scores showed superior discriminatory ability compared to ICD-AIS mapping. This is in line with previous findings reporting better performance of SRR-based compared to AIS-mapped scores in adults [30, 42-44]. Within the SRR-based methods we found no difference between the single and multiplicative ICISS, which again aligns with previously published clinical and methodical studies reporting that the most severe injury determines outcome [38, 45, 46]

Like the discrimination, model calibration was better for SRR-based than for ICD-AIS mapped scores. Hosmer-Lemeshow statistics indicated suboptimal model calibration for all scores, particularly for the MAIS and ISS. This difference may indicate a true lack of fit, but since the H-L test is sensitive to sample size, even a slight departure from a perfect fit can produce significant statistical values in very large sample sizes like ours. For that reason, we assessed the slope and intercept of calibration curves as suggested [39] and found good model calibration.

There are several limitations to our study: Model performance was tested exclusively in cases with valid ICD-AIS mapping, thereby excluding a fifth of cases and a fourth of deaths. However, we chose this approach to ensure the comparability of results between the different scoring systems, and SRRs were calculated from the complete dataset. Calibration assessment was complicated by the fact that the H-L test statistic is very sensitive to the large sample size, where minor deviations from perfect calibration can lead to statistically significant differences. To compensate for this limitation, we extracted the slopes and intercepts for calibration curves. Another important limitation that applies to all administrative datasets is the unknown accuracy of ICD coding, which could not be assessed in this study. The unsurpassable strength of this study is that all hospitalized pediatric trauma patients in the entire country were included, completely excluding a selection bias regarding the studied population.

Considering the frequent failure to derive AIS and ISS scores, the better discrimination and calibration of SRR-based scores, and the simpler calculation of SRRs compared to AIS and ISS, it seems reasonable to use SRR-based injury severity classifications for pediatric trauma research in the GHD. We assume that the high mortality in patients with a single coded injury was due to incomplete injury assessment in the most severely injured patients. This finding suggests that the single worst SRR-derived injury may be the most suitable for assessing injury severity in general pediatric trauma patients, as it is not prone to mapping failure or incomplete ICD coding. Nonetheless, ICD-AIS mapping has its role to describe injury patterns, select patients based on the injury pattern, and possible also for risk adjustment in subpopulations, e.g., patients with TBI. The results from this study reflect current injury-related mortality risk in Germany but can likely be transferred to similar health care settings.

## 5. Conclusion

This study established ICD-AIS and SRR-based injury severity classifications in the German hospital dataset to predict mortality for general pediatric trauma patients and clinically important subpopulations. All pediatric hospitalizations in Germany from 2014 to 2020 were included. Model performances were excellent regarding discrimination and calibration, with SRR-based methods outperforming ICD-AIS mapping. The results of this study provide information for researchers on appropriate methods to describe study populations control for confounding in pediatric trauma research from routine health care data, especially for research on subpopulations.

## Data Availability

The data analyzed in this study is subject to the following licenses/restrictions: the original dataset can be accessed after inquiry to the Federal Bureau of Statistics of Germany. Requests to access these datasets should be directed to https://www.forschungsdatenzentrum.de/de.

