## Appendix 1 for "Performance of ICD-10-based injury severity scores in pediatric trauma patients using the ICD-AIS map and survival rate ratios"

Supplementary materials

Appendix 1: Differences between the AIS and ISS scoring systems

| **Classification** | **AIS (Abbreviated Injury Score)** | **ISS (Injury Severity Score)** |
| --- | --- | --- |
| Anatomical regions | AIS chapters:   1. Head 2. Face 3. Neck 4. Thorax 5. Abdomen 6. Spine 7. Upper extremity 8. Lower extremity 9. External | ISS body regions   1. Head and neck 2. Chest 3. Abdominal and pelvic content 4. Extremities and pelvic girdle 5. Face 6. External |
| Scoring | Score between 1 and 6 assigned to each chapter based on the most severe injury of the chapter; score 9 for unclassifiable/unclassified injuries. | Score between 1 and 6 assigned to each body region based on the AIS classification (see left column) |
| Overall severity | Maximum AIS is calculated as the highest AIS severity score across all 9 AIS regions | ISS is calculated from the three regions with the highest scores according to the formula:  ISS= sum of the square of the 3 highest AIS  = a^2^+ b^2^ +c^2^  Important: In case one injury is 6, the overall score is set to the maximum score of 75; if one region is classified as 9, the ISS cannot be calculated |
